# Treatment of ARDS and hyperinflammation in COVID-19 with IL-6 antagonist Tocilizumab: a tertiary care experience from Pakistan

**DOI:** 10.1101/2020.06.23.20134072

**Authors:** Nosheen Nasir, Syed Faisal Mahmood, Kiren Habib, Iffat Khanum, Bushra Jamil

**Affiliations:** Assistant Professor, Dept. of Medicine, Aga Khan University, Stadium Road, Karachi 74800, Pakistan; Associate Professor, Dept. of Medicine, Aga Khan University, Stadium Road, Karachi 74800, Pakistan; Senior Instructor, Dept. of Medicine, Aga Khan University, Stadium Road, Karachi 74800, Pakistan; Professor, Dept. of Medicine, Aga Khan University, Stadium Road, Karachi 74800, Pakistan

**Keywords:** Cytokine release syndrome, Hyperinflammation, COVID-19, ARDS, IL-6, Tocilizumab

## Abstract

Cytokine release syndrome in COVID-19 is characterized by hyperinflammation which manifests as ARDS, multi-organ failure, and high inflammatory parameters. Tocilizumab, an IL-6 antagonist has been used in COVID-19 acute respiratory distress syndrome (ARDS) with conflicting results from different parts of the world. We conducted a retrospective descriptive study from Feb 2020 to May 2020 on COVID-19 patients with ARDS and hyperinflammation characterized by raised CRP and/or ferritin. A total of 244 patients with COVID-19 were admitted out of which 107 had ARDS. Thirty patients had both ARDS and hyperinflammation and received tocilizumab. The mean age was 62.5 years (SD: 13.5) and the majority were male (83%). The mean CRP pre-treatment was 217.5 mg/L and post 48 to 72 hours of tocilizumab treatment was 98.5 mg/L. Twenty-one patients (70%) also received concomitant intravenous methylprednisolone. Of the 30 patients, 7 died and 20 recovered. Ten patients required intensive care unit admission and nine developed nosocomial infections. COVID-19 associated aspergillosis was diagnosed in three patients post tocilizumab treatment. Mortality was significantly higher in patients who developed a nosocomial infection and who required intermittent positive pressure ventilation (IPPV). Our study is the first to describe the treatment outcomes with tocilizumab from a low-middle income country. The availability and cost of tocilizumab in our region which makes it imperative to understand its potential for use in our setting. Our study supports the use of tocilizumab in a select patient population with COVID-19 and recommends monitoring of nosocomial infections and opportunistic infections.

## INTRODUCTION

COVID-19 was announced as a pandemic by the World Health Organization in March 2020^1^ and since then it has infected 5,113,706 people globally and resulted in 330,361 deaths as of date^2^. The life-threatening manifestation of COVID-19 is acute respiratory distress syndrome (ARDS) and is associated with significant morbidity and mortality^3^. The underlying pathogenesis of ARDS involves a dysregulated immune response leading to a cytokine release syndrome (CRS), referring to an excessive and uncontrolled release of pro-inflammatory cytokines. CRS in COVID-19 is characterized by hyperinflammation which manifests as ARDS, multi-organ failure, and high inflammatory parameters ^4^. Key in the development of the CRS is an exaggerated release of the proinflammatory cytokine Interleukin-6 (IL-6) and elevated IL-6 levels correlate with ARDS ^5^. Marked elevation of C-Reactive Protein (CRP) (whose expression is propelled by IL-6) also serves as a biomarker to assess the severity of clinical CRS ^3^ and studies have used CRP and ferritin as surrogate markers of hyperinflammation^6,7^.

Given the pivotal role of IL-6; it has been postulated that targeting IL-6 with available IL-6 inhibitors like tocilizumab may lead to clinical suppression of the CRS ^8^. Data from clinical studies have been conflicting with regards to the efficacy of tocilizumab in COVID-19 ARDS. While limited studies from China have shown improved outcomes in COVID-19 patients with hyperinflammation and ARDS ^9^, a study from Italy did not show significant mortality benefit ^10^. While randomized controlled trials are awaited, there is an urgency to explore therapeutic options that can help to avert ICU admissions, especially given the limited capacity in resource-constrained settings. Hence, we would like to report our clinical experience of the management of ARDS and hyperinflammation with the IL-6 inhibitor Tocilizumab which will be the first from a lower-middle-income country (LMIC).

## METHODS

We conducted a retrospective descriptive study on COVID-19 patients with ARDS and hyperinflammation admitted to the Aga Khan University Hospital (AKUH); a 700-bedded tertiary care hospital. Cytokine storm or Hyperinflammation was defined as either a serum CRP ≥100 mg/L or a ferritin ≥900 ng/mLor both^6^. ARDS was defined as per WHO definition as having “onset within 1 week of a known clinical insult or new or worsening respiratory symptoms, chest imaging (radiograph, CT scan, or lung ultrasound) with bilateral opacities, not fully explained by volume overload, lobar or lung collapse, or nodules and respiratory failure not fully explained by cardiac and PaO2/Fio2 <300mmHg”^11^. Patients were excluded if they had transaminitis (ALT of greater than 3 times upper normal limit) and/or ongoing bacterial infection or tuberculosis. Demographics and clinical data from the hospital medical records were collected using a structured proforma. The outcomes assessed included in-hospital mortality, length of stay, and development of nosocomial infection during hospitalization. The study was submitted for ethical approval to the AKUH ethical review committee and received exemption (ERC reference number: 2020-3650-10382). The data was anonymized, and no personal identifiers were recorded.

Categorical variables such as gender, development of nosocomial infection were described as proportions, and continuous variables like age and length of hospital stay were described using mean, median, and interquartile range (IQR) values. Proportions for categorical variables as mentioned above were compared using the χ2 test or Fisher exact test where appropriate. Statistical analysis was performed using STATA ver 12. A p-value of less than .05 was considered statistically significant.

## RESULTS

A total of 244 patients of COVID-19 were admitted from Feb 26^th^ to May 15^th^ 2020, out of which 107 met WHO criteria for ARDS. Of these, 30 patients with ARDS who also met the criteria for hyperinflammation and qualified to receive tocilizumab. The clinical characteristics and outcomes of these 30 patients are summarized in Table 1. The mean age was 62.5 years (SD: 13.5) and the majority were male (83%). None of the patients had a rheumatological illness. The median dose of tocilizumab was 600mg (Range: 320 – 680 mg). No adverse effects were observed during or post-infusion. Twenty-one patients (70%) also received concomitant systemic steroids (intravenous methylprednisolone). Of the 30 patients, 7 died and 20 recovered while information was missing on 3 patients who left against medical advice. The mean length of hospitalization was 12 days (SD: 6.7). The mean CRP pre and post tocilizumab treatment in those who died compared to those who survived are shown in Figure 1. Ten patients required ICU admission and intermittent positive pressure ventilation (IPPV) whereas 14 patients were managed on Non-invasive ventilation (NIV). Nine patients developed nosocomial infections, of which 6 of were hospital-acquired pneumonia (three with multi-drug resistant (MDR) acinetobacter, 2 with MDR *Pseudomonas aeroginosa* and one with methicillin resistant *Staphylococcus aureus* (MRSA). Additionally, 7 patients also isolated aspergillus species from their respiratory specimens out of which 3 patients were diagnosed with COVID-19 associated aspergillosis and 4 were considered to be colonized. Mortality was higher in patients who developed a nosocomial infection (p-value: 0.005) and who required IPPV (p-value: 0.023).

**Table 1:** Clinical characteristics and outcomes of COVID-19 patients who received Tocilizumab.

| Demographics |  |  |  |
| --- | --- | --- | --- |
| Number patients (N) | 30 |  |  |
| Age mean ± SD years | 62.5 ± 13.5 |  |  |
| Gender n(%) | Male | Female |  |
|  | 25 (83%) | 5 (17%) |  |
| Length of stay mean ±SD days | 12 ±6.7 |  |  |
| Pre-treatment ventilator status |  |  |  |
| Assisted ventilation n(%) | IPPV | NIV | None |
|  | 10 (33%) | 14 (48%) | 6 (20%) |
| Drug management |  |  |  |
| Tocilizumab Dose Median (Range) mg | 600 (320-680) |  |  |
| Concomitant steroids n(%) | 21 (70%) |  |  |
| Outcome |  |  |  |
| Acute phase reactant response | Pre-tocilizumab | Post-tocilizumab |  |
| CRP mean ± SD mg/dl | 217.5± 64.6 | 98.4 ±56.3 |  |
| Immunosuppression |  |  |  |
| Nosocomial Infection n(%) | 9 (30%) |  |  |
| Aspergillus infection/colonization n(%) | 7 (24%) |  |  |
| Final outcome | Dead | Alive | Unknown |
| n (%) | 7 (23%) | 20 (66%) | 3 (10%) |

**Figure 1:**
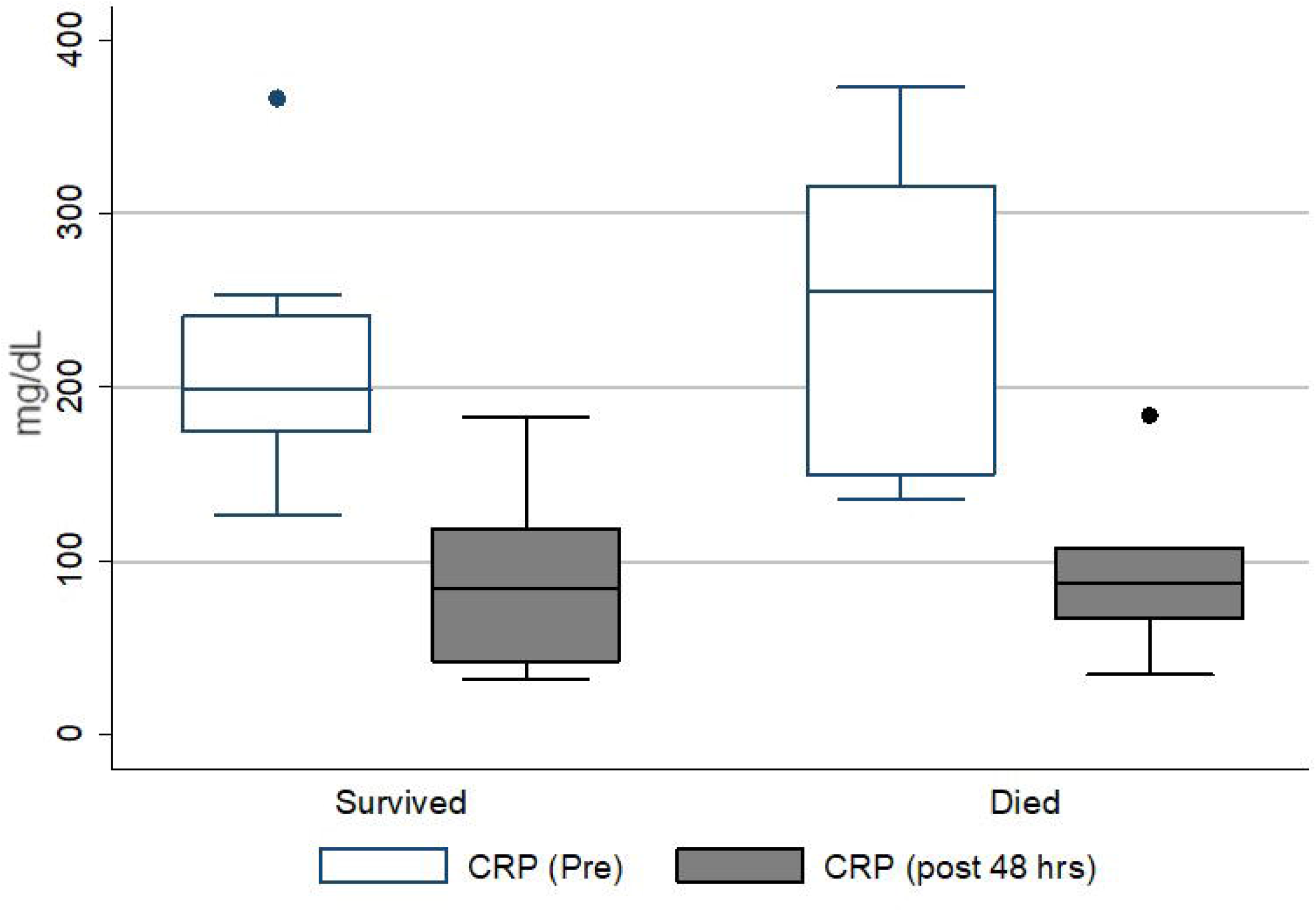
Comparison of C - reactive protein levels before and after administration of intravenous Tocilizumab in patients who died versus survived

## DISCUSSION

In a recent debriefing, WHO has raised alarm over the numbers of cases that are exponentially increasing in the lower-middle-income countries (LMICs). It is a major cause of concern particularly since the capacities of developed countries have been overwhelmed. Since the development of a vaccine is unlikely in the near future, the focus has been on treatment and compassionate use of certain medications. Tocilizumab is a monoclonal antibody targeting the receptor of IL-6; a pro-inflammatory cytokine involved in the pathogenesis of ARDS seen with COVID-19^8^. Data is urgently needed from developing countries as a “one-size fits all” strategy cannot be used in resource-constrained regions where healthcare capacities are already overstretched ^12^. Moreover, whether tocilizumab is a cost-effective option in developing countries also requires exploration because of the differences in case fatality rates from the different parts of the world suggesting that same approaches may not be regionally relevant. We conducted an observational study describing patient outcomes in those critically ill patients of COVID-19 who received tocilizumab intravenously for hyperinflammation and ARDS. Most of the data regarding its off-label use in COVID-19 has been from studies conducted in China and parts of Europe (Table 2). The age and gender distribution of our patient population were similar to those reported in studies from China and Italy ^10,13^. The mortality data has been conflicting from Italy with Colaneri M et al. reporting no benefit ^10^ whereas the Sciascia S et al.

**Table 2:** Summary of studies describing experience with Tocilizumab use in severe COVID-19 infection.

| **Study** | **Country of study** | **Number of patients who received IV TOCILIZUMAB** | **Study design** | **Outcomes** | **Conclusion** |
| --- | --- | --- | --- | --- | --- |
| Luo P et al.<sup>13</sup> | China | 15 | Observational | 3/5 (20%) mortality, 2/15 with disease aggravation | Good response in patients with Tocilizumab |
| Colaneri M et al.<sup>10</sup> | Italy | 21 | Observational | 5/21 (24%) Day-7 mortality at Day 7. No difference compared with those who didn't get Tocilizumab | Tocilizumab administration did not reduce ICU admission and mortality rate |
| Sciascia S et al.<sup>14</sup> | Italy | 34 | Pilot prospective open single-arm | 4/31 (12%) mortality in Tocilizumab arm. | May benefit in severe disease |
| Xu X et al.<sup>15</sup> | China | 21 | Observational | No mortality | Good outcome |
| Van Kraaij et al.<sup>16</sup> | Netherlands | 1 | Case report | Survived | Good outcome |
| Klopfenstein T et al.<sup>17</sup> | France | 20 | Case Control | 25% mortality in Tocilizumab group versus 48% in no Tocilizumab group | Good outcome |

showing reduced mortality in those who received treatment with tocilizumab ^14^. On the other hand outcomes have been consistently reported to be good in studies from China, at the time of submission, some had not been peer-reviewed and others did not take into account concomitant treatments ^9,13^. Our study has reported a mortality of 23% which is similar to other studies, but we also report concomitant use of systemic steroids in 70% of the patients which may have contributed in immunomodulation. None of the studies so far have reported nosocomial infection or aspergillus infection or colonization. Since IL-6 antagonism can potentially predispose to worse outcomes in infections, this is an important observation in our study and can have implications in developing countries where there is a higher incidence of multi-drug resistant infections.

Our study is limited because of small sample size and single-center design but this is expected given the availability and cost of tocilizumab in our region which makes it imperative to understand its potential for use in our setting. All the studies exploring outcomes with tocilizumab are limited due to their small sample sizes and observational design highlighting the need for randomized controlled trials. However, given the rapidity of the spread of COVID-19 infection, real-time data is needed particularly from LMICs which are about to see the peak in the number of cases.

## Conclusion

Our study supports the use of tocilizumab in a select patient population with COVID-19 and recommends monitoring of nosocomial infections and opportunistic infections in COVID-19 patients who receive the medication which may lead to adverse outcomes in these patients.

## Data Availability

Data is included within the manuscript

## SUPPLEMENTAL MATERIAL

